# Ready-To-Use Investigational Stem Cells in Patients with recent Acute Myocardial Infarction

**DOI:** 10.1101/2023.08.17.23294118

**Authors:** KC Ueng, CF Tsai, YT Chuang, JY Huang, JTK Liu

**Affiliations:** Chung Shan Medical University Hospital, Division of Cardiology, Department of Internal Medicine; Chung Shan Medical University, School of Medicine; Chung Shan Medical University, School of Medicine; HONYA Medical PTY LTD

## Abstract

**BACKGROUND:** Cardiovascular diseases are a major health concern, and stem cell therapy holds promise as a potential treatment. However, obtaining stem cells from patients who are already ill poses challenges, as does limited access to specialized laboratories. We aimed to assess the safety and preliminary effectiveness of a read-to-use investigational stem cell product, MiSaver, in patients with recent acute myocardial infarction (AMI).

**METHODS:** We enrolled 10 participants with AMI and reduced left ventricular ejection fraction (LVEF ≤45%). Group A (n=5) received a low dosage of 0.5×10^7^ cells/kg, and Group B (n=5) received a high dosage of 1.6×10^7^ cells/kg of cells. Additionally, 20 control patients who received standard care were included for statistical analysis. The primary outcome was the occurrence of side effects or graft-versus-host disease (GVHD) associated with the stem cell treatment. Secondary outcomes included changes in LVEF.

**RESULTS:** During the 12-month follow-up period, no study-related adverse effects or GVHD were observed. Baseline LVEF values for Group A and Group B were 42.0% and 41.3%, respectively, increasing to 53.0% and 53.7% at 12 months. Furthermore, no signs or symptoms of graft-versus-host disease (GVHD) were reported. Significant improvements in LVEF were observed in non-morbidly obese treated patients (cases 1-9), with baseline values of 41.3% and 12-month values of 53.7%, compared to the control group with baseline and 12-month values of 42.35% and 47.5%, respectively (p<0.05).

**CONCLUSIONS:** This trial demonstrated the safety and tolerability of MiSaver stem cells in patients with recent AMI. Preliminary evidence suggests efficacy in improving outcomes in non-morbidly obese participants.

## Introduction

Cardiovascular diseases (CVDs) are the leading global cause of death, responsible for approximately 17.9 million deaths in 2019, with myocardial infarction (MI) and stroke accounting for 85% of these fatalities [1]. Despite advancements in pharmacological and nonpharmacological interventions, MI remains a leading cause of death worldwide, accompanied by significant morbidity [2]. Consequently, there have been extensive efforts to develop alternative treatments for improving cardiac function and recovery.

Stem cell therapy has emerged as a promising option for cardiac regeneration and revascularization. Stem cells possess the ability to migrate, modulate the immune system, secrete trophic factors that enhance angiogenesis, reduce infarct volume, differentiate into mature functioning cells such as cardiomyocytes and vascular endothelial cells and improve outcomes in cardiomyopathy [4-10].

However, obtaining stem cells from patients who are already ill poses challenges, as does limited access to specialized laboratories. In light of this, our study aimed to assess the safety and preliminary effectiveness of a read-to-use investigational stem cell product, MiSaver, in patients with recent AMI. MiSaver stem cell injections, derived from umbilical cord blood, offer several advantages. They are immuno-naive and immunologically tolerant, containing various types of non-hematologic stem cells like endothelial progenitor stem cells, mesenchymal stem cells, multilineage progenitor cells, unrestricted somatic stem cells, and very small embryonic-like stem cells. Furthermore, their clinical safety has been established in allogeneic hematopoietic stem cell transplantation for more than two decades [11-17].

Multiple clinical trials have demonstrated the beneficial effects of stem cells on myocardial function in AMI patients [18-20]. The recovery of LVEF following MI is a crucial prognostic factor, with patients showing no improvement facing increased risks of sudden cardiac arrest and mortality [3]. Hence, this trial is aimed to assess the safety and effectiveness of MiSaver in promoting LVEF recovery among patients with AMI.

## Methods

### Study Design

A phase I/IIa clinical trial was conducted at Chung Shan Medical University Hospital to assess the safety and preliminary effectiveness of MiSaver stem cells in improving LVEF in patients with AMI. The stem cells utilized in the trial were provided by Honya Medical Co. Ltd. Approval was granted by the institutional review board of the clinical site, and regulatory authorization was secured from the appropriate national authority overseeing clinical trials and healthcare products.

### Participants

The trial enrolled a total of 10 patients, with 5 assigned to receive a low dose and 5 to receive a high dose of MiSaver stem cells via intravenous infusion within 3-10 days after AMI. Prior to stem cell therapy, all participants received standard treatment, including medication, percutaneous coronary intervention (PCI), and stent implantation. The enrollment of the first and second subjects in each group had a 2-week interval to allow for review by the Data Safety Monitoring Board (DSMB), with a final DSMB review conducted upon completion of both groups. Adverse events (AEs) were monitored and documented by the study team during scheduled visits and via telephone.

### Control Group

Results were compared with a retrospective control group comprising participants who received standard care (medication, PCI, and stent implantation) without stem cell therapy. The control group had a two-to-one ratio compared to the treatment group.

### Patient Eligibility

Strict eligibility criteria were applied to ensure safety and relevant results. Inclusion criteria were as follows: patients aged between 20 and 80 years, diagnosed with AMI within 7 days, elevated cardiac enzymes (CK-MB or troponin) greater than 2 times the upper limit of normal, presence of regional wall motion abnormality, and LVEF ≤ 45% on echocardiography.

Patients who were hemodynamically stable, not requiring inotropic support, with a systolic blood pressure below 80mm Hg for less than 1 hour, and a resting heart rate above 100 beats/min for less than 1 hour in the past 24 hours were eligible. Additionally, patients were required to have a peripheral artery oxygen saturation of ≥97%.

Exclusion criteria included pregnancy or breastfeeding, positive adventitious infections (such as HIV, hepatitis), need for coronary artery bypass surgery or anticipated further revascularization procedures during the 6-month study period, severe aortic or mitral valve narrowing, or evidence of life-threatening arrhythmia on baseline electrocardiogram (ECG). Patients who were unable to receive PCI examination or treatment (including NYHA Fc.IV) due to shortness of breath, had a malignant tumor, hematopoietic dysplasia, other severe organ disease, less than 1 year of life expectancy, or had chronic kidney disease with CCr<20ml/min and were on renal dialysis were also excluded from the study.

### DSMB Review

A Data Safety Monitoring Board (DSMB) consisting of a physician, a statistics professor, and a law professor was established. The DSMB conducted reviews after the initial subjects in both the low- and high-dose groups, as well as upon completion of the study. Throughout the study, any serious adverse events (SAEs) directly linked to the study product were promptly reported to the DSMB.

### MiSaver Stem Cells

MiSaver stem cells are shipped to the study site in single-use 20 ml clear vials with coated stopper seals and “flip-off” caps within two days of notification. These vials contain a heterogeneous population of stem cells derived from umbilical cord blood. Prior to shipment, each batch of vials undergoes sample testing to ensure that 80% of the cells remain viable after thawing. Additionally, the stem cells undergo comprehensive sterility testing to confirm the absence of infectious agents. This includes testing for negative culture growth to ensure sterility, negative nucleic acid testing for HIV, hepatitis C virus, and hepatitis B virus, and negative antibody testing for syphilis.

### Administrations

MiSaver stem cells are administered intravenously at a low dose of 0.5×10^7^ cells/kg (Group A, n=5) or a high dose of 1.6×10^7^ cells/kg (Group B, n=5). The stem cell vials arrive at the study site within a cryogenic container and are thawed using a temperature-controlled warmer. After thawing, the stem cell solution is diluted with a 0.9% sodium chloride solution for injection at a 1:1 ratio and used immediately. Infusion occurs over a 10-minute period at a constant and slow rate.

Patient blood type matching is important before administering MiSaver stem cells. Patients are premedicated with intravenous antihistamine (such as diphenhydramine) and corticosteroid (such as hydrocortisone) 30–60 minutes before the stem cell infusion. Any unused solution is discarded, and the total volume of injected solution takes into account the total daily fluid volume administered.

### Safety Evaluation & Parameters

Patient safety was closely monitored during hospitalization (from D0 to D3) and at scheduled follow-up visits at 1, 2, 3, 6, 9, and 12 months after treatment. Continuous monitoring of heart rate, respiratory rate, and oxygen saturation was conducted throughout the treatment period. Physiological parameters, including heart rate, blood pressure, temperature, respiratory rate, and oxygen saturation, were reviewed before infusion, at 15-minute intervals for 1 hour post-infusion, and at 30-minute intervals for 2 hours post-infusion, and any changes or adverse events were noted.

To evaluate myocardial response and reaction after stem cell therapy, serum levels of troponin and creatine kinase were tested daily from the day of admission until a noticeable decreasing trend was observed.

Follow-up visits included assessments for symptoms of graft-versus-host disease (GVHD), infection, adverse events (AEs), laboratory examinations, and ECG. Direct and indirect Coombs tests, lymphocyte surface markers reactions, PRA test-Class I and II were performed at enrollment and 3 months post-treatment, while 24-hour Holter ECG and echocardiography were conducted before treatment and at 6 and 12 months after treatment. Additionally, MRI scans were performed before treatment and at the 12-month follow-up.

Procedural complications were defined as any ventricular arrhythmia, visible thrombus formation, distal embolization, or coronary artery injury associated with the stem cell treatment procedure.

### Statistical Analysis

Quantitative data were reported as median (25th percentile -75th percentile), while qualitative data were presented as absolute frequencies and/or percentages. Fisher’s exact test was used for testing differences between therapy groups for qualitative variables due to the small sample size. The non-normal distribution of quantitative variables was identified, and the Wilcoxon signed-rank test and Wilcoxon rank-sum test were used for within-group and between-group comparisons, respectively. The stepdown Bonferroni corrected p-value was calculated for multiple comparisons. Statistical significance was defined as a two-sided p-value < 0.05. SAS 9.4 software was used for the analysis.

### Endpoints

The primary objective of this study was to assess the safety of the treatment, including the incidence of study-related adverse events (AEs) and graft-versus-host disease (GVHD). Secondary endpoints included evaluating the change in LVEF using echocardiography at admission, as well as at 6 and 12 months post-treatment. Functional assessment was also performed using the New York Heart Association (NYHA) and Canadian Cardiovascular Society (CCS) classification systems.

## Results

### Baseline Characteristics

Ten male adult patients (median 55.4, range: 41-70 years) were enrolled in the MiSaver study between January 2021 and March 2022. Most participants had risk factors for MI, including hypertension, hyperlipidemia, diabetes mellitus, and a history of smoking (Figure 1). The control group (Group C) consisted of 20 patients diagnosed with AMI and LVEF < 45% at admission, who received standard intervention and had available LVEF echocardiogram follow-up results from 3 to 12 months at the trial hospital. There were no significant differences in age or baseline LVEF between the two groups. Of the 20 patients in the control group, 17 were male and 3 were female.

**Figure 1.**
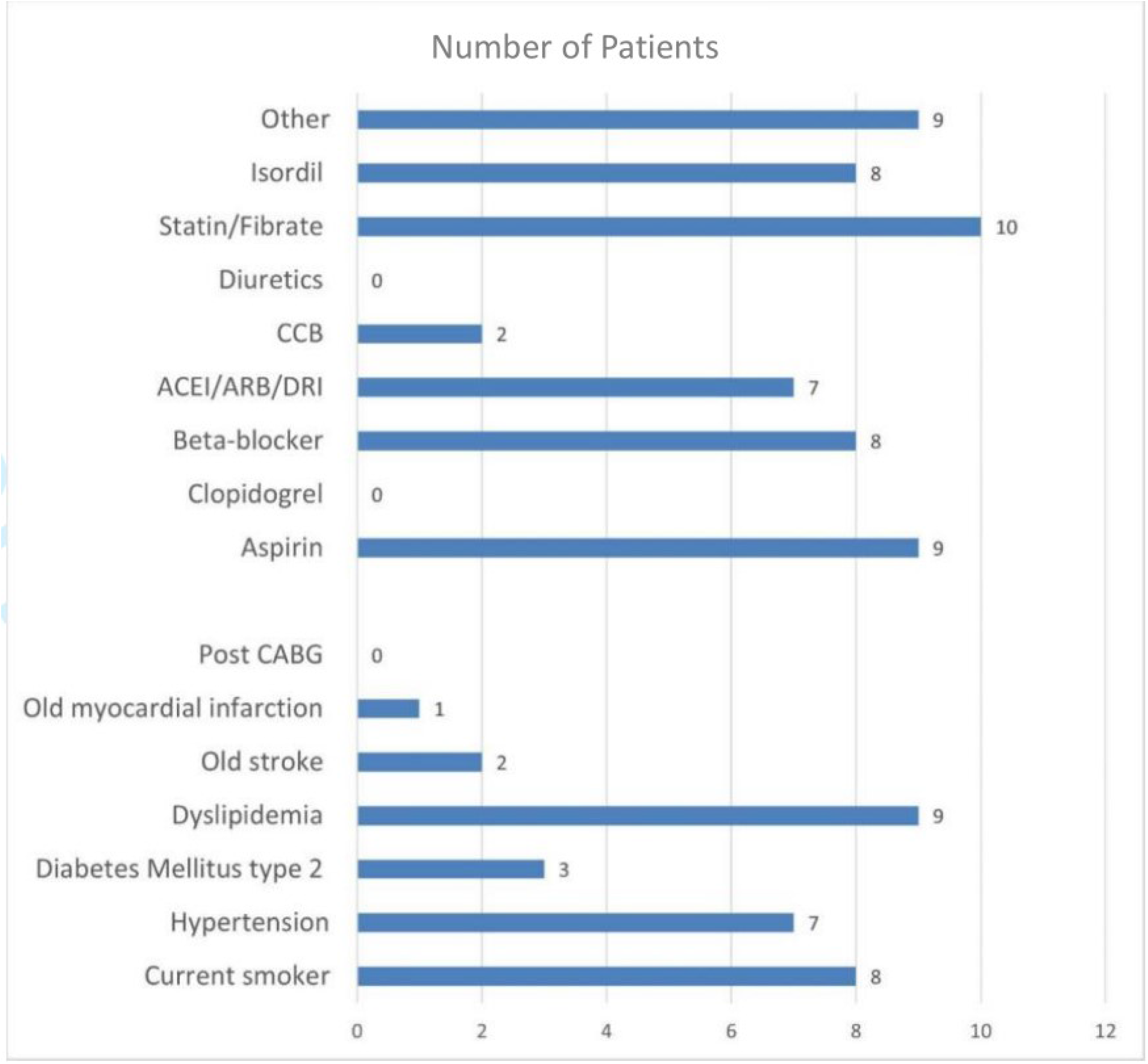
Baseline characteristics of participants

### MiSaver Infusion

Baseline clinical laboratory tests, echocardiography, MRI, and 24-hour Holter EKG were obtained before the stem cell infusion. Participants received either a dose of 0.5 or 1.6 × 10^7^/kg of ABO/Rh matched MiSaver stem cells intravenously within 2-5 days post-MI (Table 1).

**Table 1.** Baseline characteristics of participants age, weight and MiSaver infusion time and dosages.

| Participant | Age(range) | Infusion,<br>days post MI | Body weight range<br>(kg) | MiSaver Cell Dose<br>x107/kg |
| --- | --- | --- | --- | --- |
| 1 | 66-70 | 4 | 66-70 | 0.5 |
| 2 | 41-45 | 2 | 76-80 | 0.5 |
| 3 | 56-60 | 3 | 61-65 | 0.5 |
| 4 | 51-55 | 3 | 56-60 | 0.5 |
| 5 | 61-65 | 2 | 76-80 | 0.5 |
| 6 | 66-70 | 4 | 56-60 | 1.6 |
| 7 | 41-45 | 2 | 96-100 | 1.6 |
| 8 | 66-70 | 5 | 66-70 | 1.6 |
| 9 | 41-45 | 3 | 71-75 | 1.6 |
| 10 | 41-45 | 3 | 96-100 | 1.6 |
| Average |  | 3.1 (2~5) | 74.3 (56.8~100) | 1.1 (0.5~1.6) |

### Safety

The primary objective of the trial was to comprehensively assess the safety of MiSaver stem cells. Over the course of the 12-month follow-up period, no instances of mortality, recurrent MI, or adverse events directly related to the study were observed. Furthermore, no signs or symptoms of graft-versus-host disease (GVHD) were reported. These findings indicate that the administration of MiSaver stem cells was well-tolerated and did not pose any significant safety concerns.

### Recovery of LVEF on Echocardiogram

The preliminary analysis compared Cases 1 to 9 with the control group (C) due to a delay caused by the COVID-19 pandemic before enrolling Case 10.

There was no statistically significant difference of LVEF at admission between the treatment groups (Cases 1-9) and the control group (A+B: 41.3 (38.3-42.3), C: 42.35 (32.7-45), p=0.6459); however, at the 12-month follow-up, LVEF of the treatment groups (Cases 1-9) demonstrated a statistically significant improvement in LVEF compared to the control group, with measurements of 53.7% (46.1-56.1) versus 47.5% (39.35-53.4) respectively. (p=0.0455) (Table 2).

**Table 2.**
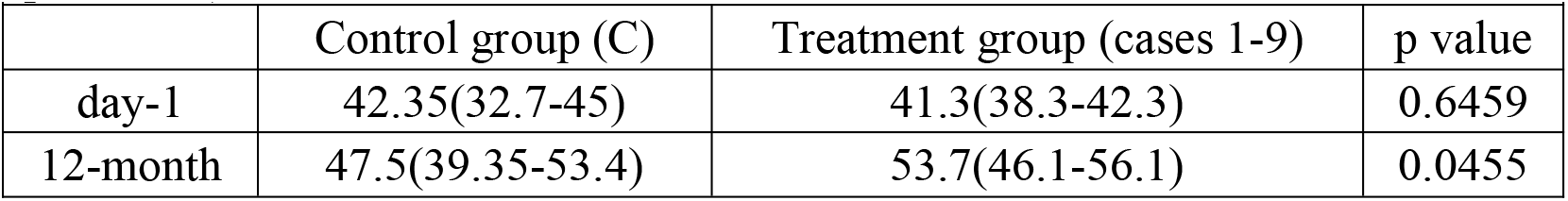
Left Ventricular Ejection Fraction. Cases 1-9. Median (25th -75th percentile)

| Table 2. Left Ventricular Ejection Fraction. Cases 1-9. Median (25th - 75th percentile) |  |  |  |
| --- | --- | --- | --- |
|  | Control group (C) | Treatment group (cases 1-9) | p value |
| day-1 | 42.35(32.7-45) | 41.3(38.3-42.3) | 0.6459 |
| 12-month | 47.5(39.35-53.4) | 53.7(46.1-56.1) | 0.0455 |

Upon completion of the follow-up for Case 10, the treatment groups (A+B or Cases 1-10) showed an improvement in LVEF of 53.4% (42.6-56.1), representing a median increase of 11.7% compared to a 5.1% improvement in the control group. However, the difference between the groups was not statistically significant at baseline or the 12-month follow-up (Figure 2)(Table 3).

**Table 3.** Left Ventricular Ejection Fraction (%). Median (25th -75th percentile)

| Table 3. Left Ventricular Ejection Fraction (%) . Median (25th - 75th percentile) |  |  |  |
| --- | --- | --- | --- |
|  | Control group (C) | Treatment group (cases 1-10, A+B) | p value |
| day-1 | 42.4 (32.7-45.0) | 41.7 (38.3-42.6) | 0.8029 |
| 12-month | 47.5 (39.4-53.4) | 53.4 (44.8-56.1) | 0.1518 |
| ΔMedian LVEF | 5.1% | 11.7% |  |

**Figure 2.**
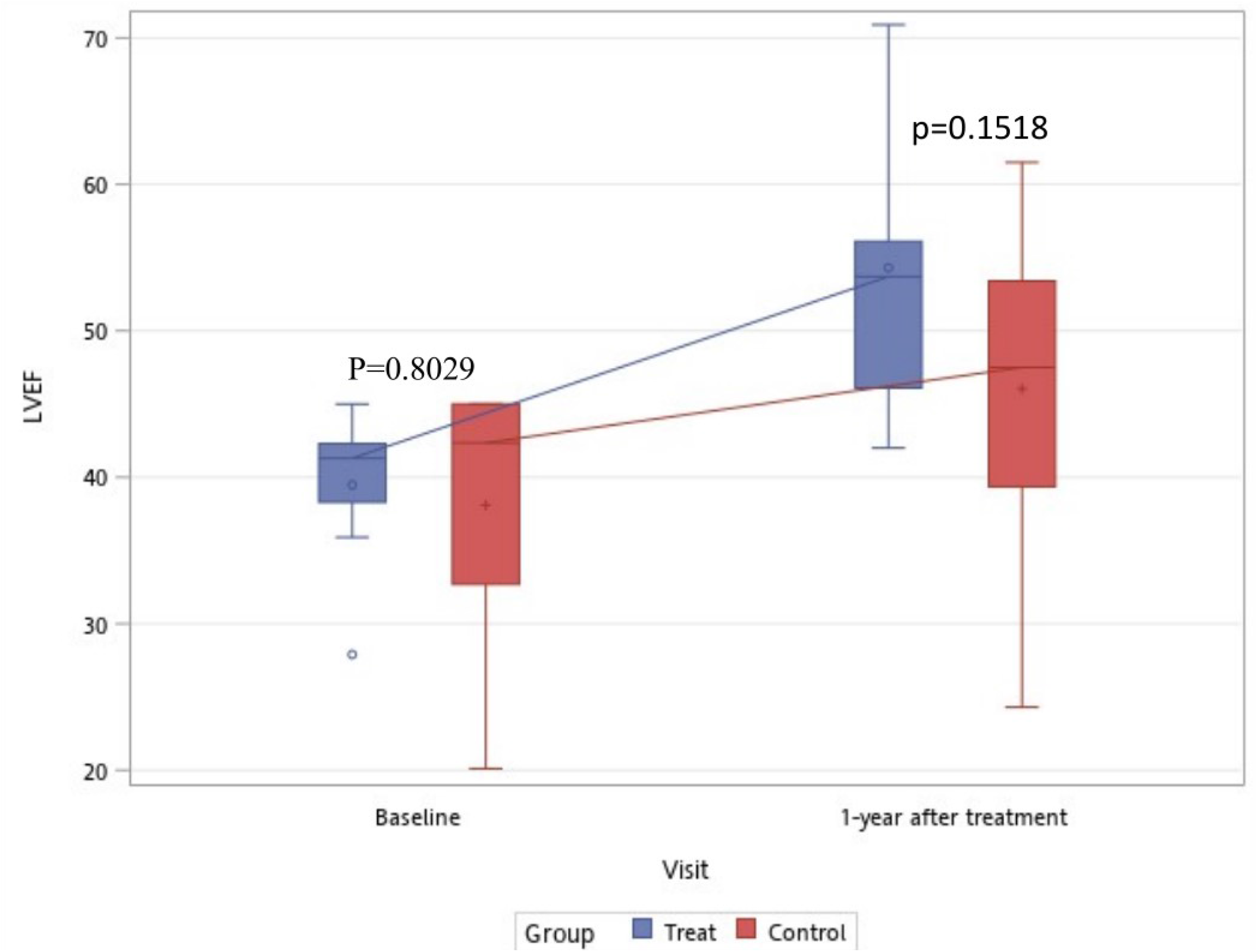
LVEF of treatment group (A+B) and control group C at base line and 12 months.

Both the low-dose (A) and high-dose (B) treatment groups exhibited an increase in LVEF from baseline (A: 42 (38.3-42.3), B: 40.65 (33.95-41.95)) at 6 months (A: 49.6% (48.3-52.1), B: 53 (47.5-61.8)) and 12 months (A: 53% (46.1-68.6), B: 54.7 (48.15-55.9)) (Figure 2). No significant differences were observed between the two groups at baseline, 6 months, or 12 months (p=0.4567, 0.3886, or 0.5871, respectively) (Figure 3). Additionally, there was no statistically significant difference in improvement from 6 months to 12 months for the combined treatment groups (A+B) (Table 4) (Figure 4).

**Table 4.** LVEF of treatment group (A+B) at base line, 6 months and 12 months. Median (25th percentile -75th percentile)

|  | Treatment group |
| --- | --- |
| day-1 | 41.7 (38.3-42.6) |
| 6-month | 50.4 (43.8-54.8) |
| 12-month | 53.4 (44.8-56.1) |
| Paired t-test, p value |  |
| baseline vs 6-month | 0.0012 |
| baseline vs 12-month | 0.0012 |
| 6-month vs 12-month | 0.6160 |

**Figure 3.**
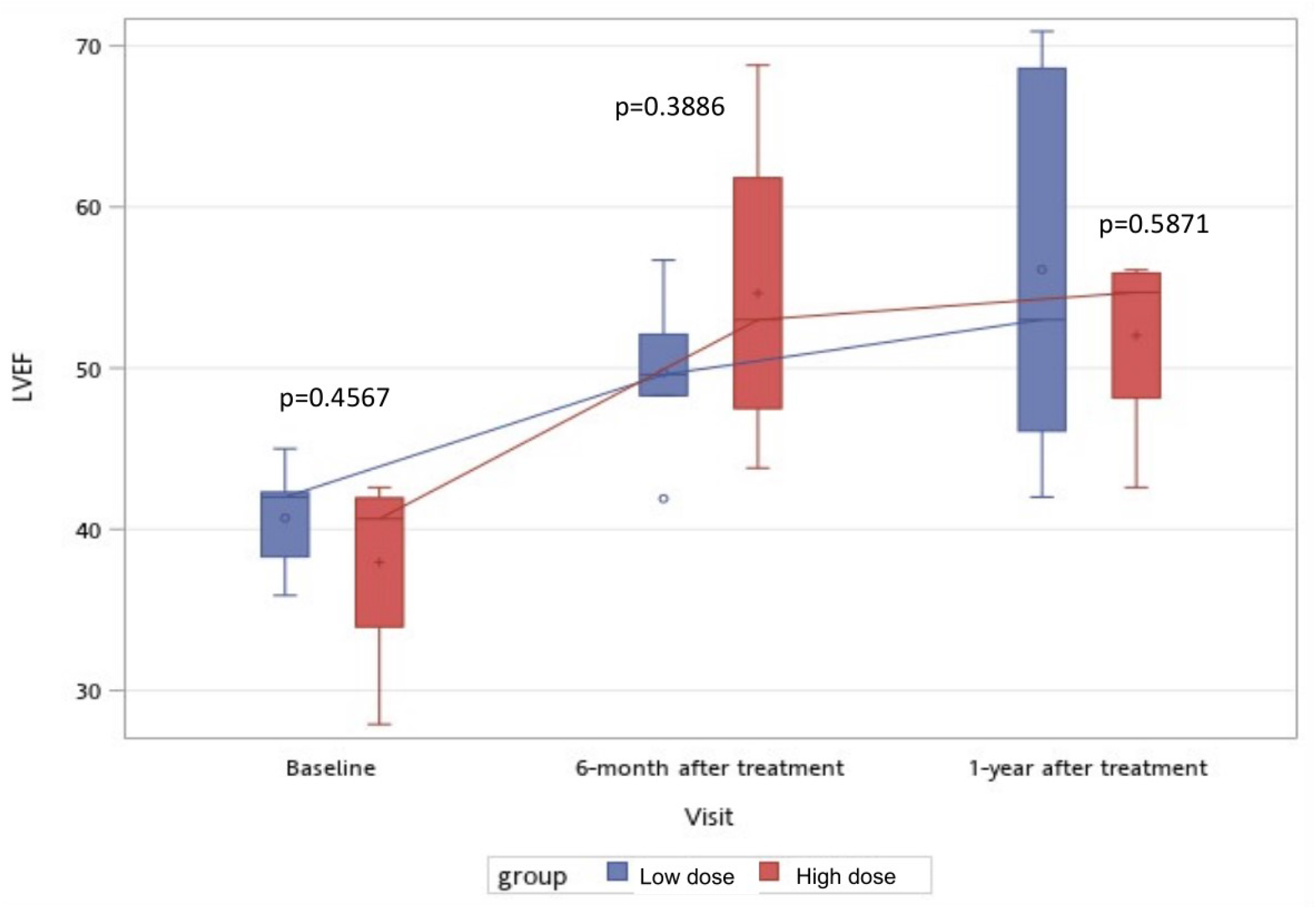
Graph shows the mean LVEF values at baseline, 6 months, and 12 months for low (A) and high (B) dosage groups. Both groups experienced an increase in LVEF from baseline (A: 42 (38.3-42.3), B: 40.65 (33.95-41.95)) to 6 months (A: 49.6% (48.3-52.1), B: 53 (47.5-61.8)) and to 12 months (A: 53% (46.1-68.6), B: 54.7 (48.15-55.9)). There was no statistical difference between the two groups at baseline, 6 months, or 12 months(p = 0.4567, 0.3886, or 0.5871, respectively).

**Figure 4.**
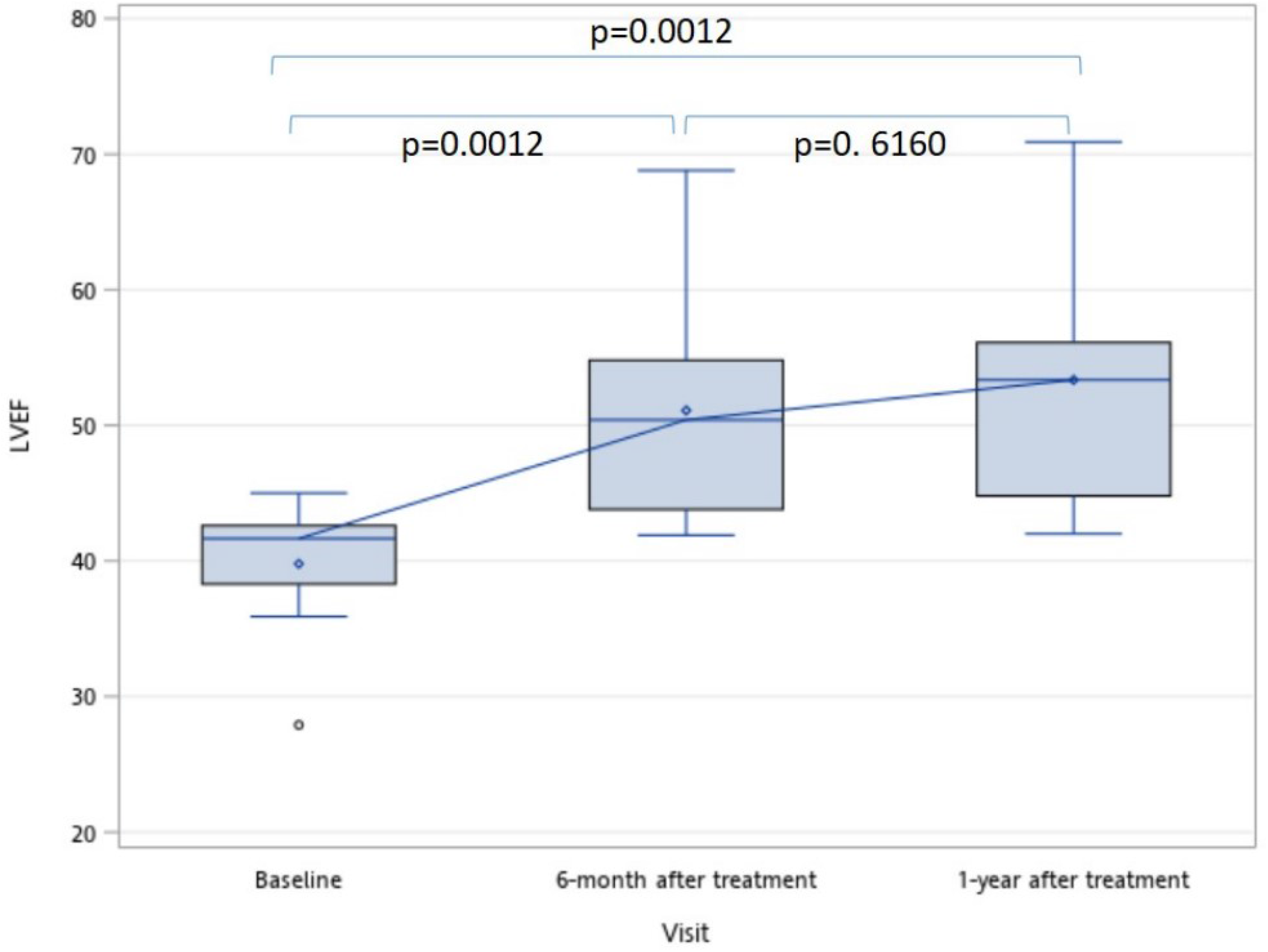
LVEF of treatment group (A+B) at base line, 6 months and 12 months.

### Functional Classification Outcomes

Functional improvement was assessed using both the NYHA and CCS classification systems at baseline, 6 months, and 12 months after treatment.

All participants showed improvement in their CCS functional activities, with an increase in their classification score. At baseline, seven participants had a CCS classification of II and three participants had a CCS classification of I (Table 5). Over the 12-month period, six participants (60%) had one functional class improvement, and four had two functional class improvements.

**Table 5.** Functional Classification of Angina. CSS, Canadian Cardiovascular Society. NYHA, New York Heart Association.

|  | CCS |  |  | NYHA |  |  |
| --- | --- | --- | --- | --- | --- | --- |
| Participant | Baseline | 6 months | 12 months | Baseline | 6 months | 12 months |
| 1 | 1 | 1 | 0 | 1 | 1 | 1 |
| 2 | 2 | 1 | 1 | 2 | 1 | 1 |
| 3 | 2 | 1 | 1 | 2 | 1 | 1 |
| 4 | 2 | 0 | 0 | 2 | 1 | 0 |
| 5 | 1 | 0 | 0 | 2 | 1 | 1 |
| 6 | 2 | 1 | 0 | 2 | 1 | 1 |
| 7 | 2 | 1 | 0 | 2 | 1 | 1 |
| 8 | 2 | 1 | 1 | 2 | 2 | 2 |
| 9 | 1 | 1 | 0 | 2 | 1 | 1 |
| 10 | 2 | 0 | 0 | 2 | 1 | 1 |
| Average | 1.7 | 0.7 | 0.3 | 1.9 | 1.1 | 1.0 |

At baseline, nine participants had a NYHA Classification of II, and one participant had a NYHA Classification of I (Table 5). Over the 12-month period, seven participants experienced one functional class improvement, and one participant experienced two functional class improvements.

## Discussion

This clinical trial assessed the safety of a read-to-use, investigational stem cell product, providing access to stem cells without the need for laboratory facilities or laboratory work. The trial also aimed to test its preliminary efficacy as a secondary objective.

During the 12-month follow-up period, all participants survived, and there were no instances of mortality, recurrence of MI, or adverse events directly related to the study. Additionally, no signs or symptoms of graft-versus-host disease were reported.

Both the low-dose (Group A) and high-dose (Group B) groups exhibited an increase in LVEF at 6 months and 12 months. The high-dose group showed a median 3.4% greater improvement in LVEF compared to the low-dose group at 6 months, suggesting a possible earlier recovery of left ventricular function. However, due to the small study group size, statistical significance was not found, and further investigation in larger clinical trials is required.

The LVEF of the combined treatment groups (A+B) demonstrated a notable medium improvement of 11.7 % throughout the study period, increasing from a baseline of 41.7% (38.3-42.6) to 53.4% (44.8-56.1) at 12 months. However, upon including case 10 in the analysis, the previously observed statistical significance related to the improvement of LVEF was no longer present. To understand this finding, we reviewed case 10 and compared it to the other participants. We observed the least improvement in LVEF over the 12 months. Notably, case 10 had the highest BMI among all participants and falls under the classification of morbid obesity.

Obesity is known to be a significant cardiovascular and health risk, and it can potentially hinder the recovery process in individuals who have experienced a heart attack. It is also associated with an increased risk of complications, including heart failure, arrhythmias, infections, and delayed wound healing. Moreover, obesity may reduce the effectiveness of treatments and interventions.

To ensure more accurate and conclusive findings regarding the improvement of LVEF with stem cell treatment after AMI, we suggest future trials consider high BMI as an exclusion criterion. This approach would help isolate the effects of interventions on individuals without the confounding influence of obesity.

All participants in the study exhibited improvement in the functional classification scale. However, to determine the significance of this improvement and accurately assess the specific impact of stem cells on the observed outcomes, further investigation with a controlled placebo group is recommended.

Although renal function was closely monitored, the limited data from the control group were insufficient for a statistical analysis. Nonetheless, the observations suggest a potential protective or therapeutic trend, indicating the need for further investigation through larger-scale trials or independent renal trials.

Based on the safety profile, preliminary efficacy, and ease of use of MiSaver stem cells, further larger-scale phase IIb/III clinical trials are necessary to test their therapeutic effects in cardiomyopathic disorders. Establishing appropriate distribution facilities will ensure easy accessibility to these off-the-shelf stem cells and facilitate testing of their efficacy.

## Conclusion

The results of this phase I/IIa trial provide evidence of the safety and tolerability of intravenous infusion of MiSaver stem cells in patients with AMI. Statistically significant improvements in LVEF were observed in non-morbidly obese participants, suggesting the potential therapeutic benefit in improving cardiac function. However, further investigation is needed to explore any potential differences and their significance in larger clinical trials. These findings support the ongoing development and investigation of MiSaver stem cells as a potential therapy for AMI.

## Data Availability

All data produced in the present study are available upon reasonable request to the authors.

## Limitations

This study has several limitations, which may affect the interpretation of the results. Firstly, due to ethical considerations, no patients received a placebo, and the control group was based on retrievable hospital records with equivalent enrollment criteria. Standard hospital care was supported by the governmental public health system, and limited laboratory and examination results were available for significant statistical analysis. Secondly, only male patients were enrolled in the study, despite both sexes being eligible to participate. This may limit the generalizability of the findings to the broader population. Thirdly, due to the COVID-19 pandemic during the trial period, a limited number of patients were enrolled using pre-specified strict inclusion/exclusion criteria. Participants were subject to quarantine during the study period, which may have influenced the exact follow-up time or results. However, despite the pandemic, all participants survived, indicating the safety of the intervention. Future studies should aim to address these limitations to improve the generalizability and validity of the findings.

## Contributorship

Dr. Ueng, Dr. Tsai, and Dr. Chuang served as Project Investigators for the clinical trial. Huang conducted the data analysis. Dr. Ueng and Dr. Liu were responsible for the clinical design and writing of the manuscript.

## Acknowledgments and Special Thanks

We extend our special thanks to Chung Shan University Hospital for executing the clinical trial and providing sponsorship (Research Project: CSH-2018-C-005). We also acknowledge HONYA Medical for sponsoring MiSaver stem cells.

## Funding, Grant/Award Info

This clinical trial is co-funded by Chung Shan University Hospital (Research Project: CSH-2018-C-005) and HONYA Medical.

## Data Sharing Statement

The data collected during this study will be made available to academic organizations upon request. However, access to the data is subject to certain conditions and restrictions to ensure participant confidentiality and privacy. Interested parties may contact the corresponding author for further details.

## Ethical Approval and Compliance with the Declaration of Helsinki

This clinical trial was conducted in accordance with the ethical principles outlined in the Declaration of Helsinki. The research protocol received approval from the Institutional Review Board of Chung Shan Medical University Hospital, reference numbers CS19037 and CS22019. Informed consent was obtained from all participants (or their legal guardians), ensuring the protection of their rights, welfare, and privacy.

